# A qualitative study on cultural determinants for breastfeeding cessation and their association with child diarrhoea in the rural Peruvian Andes

**DOI:** 10.1101/2021.02.25.21252475

**Authors:** Néstor Nuño Martínez, Jordyn Wallenborn, Daniel Mäusezahl, Stella M. Hartinger, Joan Muela Ribera

**Affiliations:** Swiss Tropical and Public Health Institute. Socinstrasse 57, P.O.Box CH-4002, Basel, Switzerland; University of Basel. Petersplatz 1, P.O. Box CH-4001, Basel, Switzerland; Universidad Peruana Cayetano Heredia. Av. Honorio Delgado 430, urb. Ingeniería, S.M.P., Lima, Peru; Partners for Applied Social Sciences (Pass-International), Baal 58, 3980 Tessenderlo, Belgium; Universitat Rovira i Virgili, Avinguda Catalunya 35, 43005 Tarragona, Spain

**Keywords:** Breastfeeding, child, child diarrhoea, cultural factors, female, local explanatory models, peer-counselling, pregnancy, social norms, Peru

## Abstract

**Background:** In some parts of the world, breast milk is seen as a potential source of child diarrhoea. While this belief has been explored in African and Southeast Asian countries, it remains vastly understudied in Latin American contexts. We investigate cultural factors contributing to breastfeeding cessation in rural high-altitude populations of the Peruvian Andes. The role of cultural factors in the local explanatory model of child diarrhoea, and whether these perceptions are integrated in the local healthcare system were assessed.

**Methods:** We carried out semi-structured interviews with mothers (n=40) from 29 communities participating in a large community-randomised controlled trial and health personnel (n=15). We used trial’s baseline information to describe characteristics of women participants.

**Results:** Cultural beliefs for breastfeeding cessation included the perception that breast milk turned into “blood” after six months and that breastfeeding caused child diarrhoea. We identified eight local types of child diarrhoea, and women associated six of them with breastfeeding practices. “Infection” was the only diarrhoea mothers linked to hygiene and the germ disease concept and perceived as treatable through drug therapy. Women believed that other types of diarrhoea could not be treated within the formal healthcare sector. Interviews with health personnel revealed no protocol for, or consensus about, the integration of the local explanatory model for child diarrhoea in local healthcare and service provision.

**Conclusion:** The local explanatory model in rural Andean Peru associated breastfeeding with child diarrhoeas. Cultural beliefs regarding diarrhoea management may increase home treatments, even in cases of severe diarrhoeal episodes. Future breastfeeding interventions should promote peer-counselling approaches to reduce negative attitudes towards breastfeeding and health practitioners. Local explanatory models should be incorporated into provincial and regional strategies for child diarrhoea management to promote equity in health and improve provider-patient relationships.

## Background

Diarrhoea is one of the major causes of morbidity and mortality in children under five years [1]. Recurrent diarrhoea episodes can produce malnutrition and increase risk of other serious diseases such as respiratory infections, measles, and pneumonia [2-5]. The occurrence of diarrhoea and malnutrition at young ages can lead to significant long-term adverse effects in adulthood, such as non-communicable disease or reduced economic productivity [6-8].

Local concepts, perceptions on causes and treatment options and practices in dealing with child diarrhoea exist cross-culturally and are generally referred to as local explanatory models [9-10]. Many of these local types of child diarrhoea are not commonly linked to hygiene and the germ disease concept, as they originate from environmental or supernatural events [10-12]. In addition, they are typically present and most widely accepted in locations where education levels are low, healthcare systems are weak and traditional medicine or elder family/community members are respected sources of information [13-16]. Local beliefs and concepts regarding causes of child diarrhoea are complex and, in some cases, even contradictory. For example, in Tanzania some ethnic groups consider diarrhoea a life-threatening ailment while in Nigeria or Thailand other groups seen it as a normal sign of growth and development [16, 17, 18]. These perceptions also include interpretations regarding diarrhoea management practices. In some parts of rural Vietnam, mothers consider breastfeeding beneficial during diarrhoea episodes [15]. On the other hand, in rural Thailand, mothers believe that breastfeeding during diarrhoea episodes worsen the illness [16]. Similarly, in Pakistan India, and Sri Lanka, some ethnic groups believe that breast milk can be contaminated through witchcraft, a new pregnancy, the mother’s illness, or exposure to environmental factors [19, 20].

Breastfeeding is recognised as a cost-effective practice that prevents infections and diarrhoea during infancy, and reduces risk of dehydration, severe disease progression and disease duration [21-24]. In fact, estimates suggest that an increase in breastfeeding levels worldwide would prevent almost one million annual deaths in children [8]. The World Health Organization (WHO) recommends exclusive breastfeeding for the first six months of life, followed by complementary breastfeeding until the age of two [25]. Despite the well-known benefits of breastfeeding, about 63% of children under six months of age are not exclusively breastfed in low- and middle-income countries (LMIC), and rates of breastfeeding cessation increase exponentially after six months of age [8]. The majority of research to date investigates demographic factors associated with breastfeeding cessation [26-30]; however, cultural beliefs may also influence breastfeeding.

Different studies around the world examined the associations between cultural reasons for breastfeeding cessation and local explanatory models of child diarrhoea, but little is known about these perceptions in the rural Peruvian Andes. Despite being a middle-income country, Peru is characterised by large social and health disparities. Forty-eight percent of all children in the lowest poverty quintile suffer from malnutrition compared to 5% in the richest quintile [31]. According to the 2014 National Demographic and Family Survey, Peru has a high reported two-week prevalence of diarrhoea (12%) in children <5 years of age. In addition, 68% of children <6 months were exclusively breastfeed, with a median duration of 4.6 months. Levels of exclusive breastfeeding and breastfeeding duration were higher in rural areas (83% and 5.1 months) and among mothers with lower education [32]. Previous studies conducted in high-altitude Andean settings show that breastfeeding practices follow culture-specific beliefs. This implies that mothers may cease exclusive breastfeeding temporarily or permanently according to local illness explanatory models [33-36]. Considering the impacts of cultural beliefs on breastfeeding practices and child diarrhoea management in Andean settings, our study: i) identifies cultural factors for breastfeeding cessation and their association with the local explanatory model of child diarrhoea in rural high-altitude Andean Peru and, ii) explores whether these perceptions and beliefs are integrated in the local healthcare and service provision.

## Methods

### Study site and participants

Women participants were identified from a community-randomised controlled trial (henceforth referred to as the “IHIP-2” trial [37]) evaluating the impact of health and early child development home-based interventions in 82 rural communities. IHIP-2 took place in San Marcos and Cajabamba, two rural high-altitude resource-limited provinces situated in Northern Andean Peru (Cajamarca). IHIP-2 was conducted among households with children <18 months that were not enrolled in the national early child development (ECD) programme and belonged to one of the four trial arms with either i) an improved biomass cookstove, or ii) an ECD intervention, iii) both interventions or iv) none (control). The majority of families participating in IHIP-2 lived in adobe households and practised subsistence agriculture (i.e. agriculture outputs are used to meet familial needs). Breastfeeding counselling in the setting was mostly provided through health personnel at healthcare centres. A full description of the site, participants and IHIP-2 design is found in Hartinger *et al*. [37].

### Study design

The study complied with the guidelines of the Consolidated Criteria for Reporting Qualitative Research (COREQ) [38]. Our qualitative study was conducted between June and December 2016. Forty women (10 per trial arm) from 29 of the trial communities were enrolled purposively based on their positive attitudes with IHIP-2 fieldworkers. Sample size was determined by convenience on the basis of the logistic and human resources available. We also contacted local health personnel working in six healthcare centres in the region applying the sample-referral sampling technique. Sample size for assessing health personnel was determined by saturation, i.e., recruitment stopped when no new relevant data emerged on the research topic [39]. All study participants were approached face-to-face. Two health practitioners refuse to participate due to limited time availability. Semi-structured in-depth interviews with women participants were carried out in their households to identify factors regarding breastfeeding cessation and investigate their association with the local explanatory model of child diarrhoea. Interview guides were based on previous studies that qualitatively explored the associations between cultural factors for breastfeeding cessation and child diarrhoea [17, 40]. We also conducted semi-structured interviews with health personnel in health centres to assess integration of women perceptions and beliefs within the local healthcare and service provision. We designed interview guides for health personnel using information analysed from interviews with women participants. Interview guides for women and health personnel were piloted with two participants from each group. No repeated interviews were conducted. All interviews were carried out in Spanish, tape-recorded, and had an approximate duration of one hour. No extra field notes were taken during interviews. All interviews were conducted by an anthropologist (NN) specialised in Medical Anthropology and Sociology. At the time of the study, NN was part of IHIP-2 coordination team. This characteristic was reported to all study participants. NN established a previous relationship with women participants (informal contacts during IHIP-2 home-visits) prior to study commencement. Women were informed that the research sought to understand childhood diseases in the area according to their knowledge and experiences. No previous contact was made with health personnel. For them, the main study goal presented was to understand their knowledge, attitudes and practices about local perceptions of child disease. No other person besides the study participants and NN were present during the interviews.

### Data analysis

Interviews were transcribed verbatim by NN using Express Scribe transcription software (NCH software, Canberra, Australia). Transcripts were not returned to participants for comments or corrections. NN then conducted a manual standard content/thematic analysis [39]. First, information was read, edited, derived in basic themes and organised into categories according to the designated analytical guides (e.g., factors for breastfeeding cessation, local types of child diarrhoea). Secondly, NN and JM compared information among categories to identify different opinions among participants. Finally, a general interpretation for each category was derived from the comparative analysis. NN and JM selected appropriate quotes to illustrate findings. Any new themes that emerged during the process were included in the analysis. A description of the transcriptions and coding tree can be provided upon request. Study participants did not provide feedback on the findings. Baseline information from IHIP-2 was used to describe characteristics of women participants. Data were entered in Census and Survey Processing System 6.3 (United States Census Bureau, USA) and then exported to STATA 15.0 Statistical software (STATA CORP. College Station, Texas, USA) for analysis. We performed a descriptive analysis using means and percentages [37].

## Results

### Women characteristics and changes in breastfeeding practices

The majority of women participants lived in households with adobe walls, earthen floors, tiled roofs, and electricity. Households had an average of 2.5 rooms and the mean number of inhabitants per household was 4.7. Women were mostly married or had a civil partnership (85%), and their mean age was 28 years. Women average years of schooling was 6.0 and the primary and secondary occupations among them were housework (97%) and self-employment (55%). On average, they had 2.2 children. At the time of the study, mean age of children participating in IHIP-2 was 1.8 years and 55% of women (N=22) had ceased breastfeeding. Among those mothers who ceased breastfeeding, 85% (N=18) declared having done so before their child reached two years of age. Households were balanced across the different quintiles of poverty (Table 1).

**Table 1.** Household, demographic, and wealth characteristics of women participants and their children

|  | N | N (%) or Mean (SD) |
| --- | --- | --- |
| *Household characteristics* | 40 |  |
| Adobe wall type |  | 36 (90.0) |
| Earthen floor type |  | 35 (87.5) |
| Roof tile type |  | 33 (82.5) |
| Electricity |  | 31 (77.5) |
| Number of rooms |  | 2.5 (1.0) |
| *Demographic characteristics* | 40 |  |
| Number of inhabitants per household |  | 4.7 (1.4) |
| Civil status (married/civil partnership) |  | 34 (85.0) |
| Maternal age (years) |  | 28.1 (8.1) |
| Maternal years of schooling<sup>1</sup> |  | 6.0 (3.8) |
| Primary maternal occupation (housework) |  | 39 (97.5) |
| Secondary maternal occupation (self-employed) |  | 22 (55.0) |
| Number of children per woman |  | 2.2 (1.3) |
| Child age (years)<sup>2</sup> |  | 1.8 (0.5) |
| Breastfeeding cessation<sup>2</sup> |  | 22 (55.0) |
| Before the child reached two years | 21 | 18 (85.7) |
| *Wealth classification<sup>3</sup>* | 40 |  |
| 1. Quintile (lowest) |  | 8 (20.0) |
| 2. Quintile |  | 8 (20.0) |
| 3. Quintile |  | 8 (20.0) |
| 4. Quintile |  | 9 (22.5) |
| 5. Quintile (highest) |  | 7 (17.5) |

<sup>1</sup> Includes primary (1-6 years) and secondary (7-12 years) education. For higher degrees, we considered: higher (not university) education not completed (12.5 years), higher (not-university) education completed (14 years), university education not completed (13.5 years) and university education completed (16 years).
<sup>2</sup> Information collected at the time of study, six months after IHIP-2 baseline. It refers to mothers/children participating in IHIP-2.
<sup>3</sup> Calculated using the nationally validated Young Lives wealth index [41].

### Factors for breastfeeding cessation

All women reported receiving regular breastfeeding counselling in local healthcare centres. In general, they indicated that breastfeeding helped children to grow up stronger and develop their cognitive potential. They mentioned four major reasons for breastfeeding cessation based on biological, social, and cultural factors. The first reason was low breast milk supply. Women believed that this was caused by a high number of pregnancies within a short time period. Time commitment was mentioned as another reason for breastfeeding cessation. Women declared that continuous children’s demands to breastfeeding made it difficult to complete daily chores, leading to distress and fatigue. Thirdly, some women believed that the quality of breast milk started declining sixth months after childbirth, and then loses its nutritional value and becomes a drink made of blood: “Breastfeeding is good until the sixth month, but never after. This is because breast milk becomes *chuvia* [*soft*], like water. It is no longer dense” (Interview with Ana, September 20, 2016). Some of these women associated changes in the structure and composition of breast milk with their own diet. A diet rich in legumes and animal protein would lead to good-quality breast milk. In contrast, excessive consumption of cereals, rice, and potatoes would make the breast milk *chuvia*. In general, women found that breastfeeding represented a communication channel between mothers and children. This perception had beneficial connotations such as the transmission of positive emotions and the strengthening of mother-child bonds. However, mothers also considered that breast milk could cause child diarrhoea, and was cited as another reason for breastfeeding cessation. The relationship between breastfeeding and child diarrhoea is described in the following section:

*“Breastfeeding was not good for her* [my child]. *She was always ill. She seemed to recover but then it* [the disease] *came again. People told me to cease breastfeeding, that it was the problem [for her repeated illnesses]. People also explained to me that if adults are ill then the disease is transmitted to children through breastfeeding* […] *Breast milk can produce many types of diarrhoeas”* (Interview with Laura, July 7, 2016).

### Breastfeeding and the local explanatory model for child diarrhoea

Overall, women participants indicated that breastfeeding was a potential cause of child diarrhoea when it was carried out within certain contexts and periods:

*“Children get everything from breast milk. When they are sick, we* [mothers] *have to take the medicine too to help them recover* […] *Even if mothers are sick, children can get sick from the breast milk* […] *In addition, what mothers eat regularly can also affect children’s health if they breastfeed” (Interview with Marina, August 8, 2016)*.

According to mothers, child diarrhoea in the local illness model was caused by either i) “hot milk” (*leche soledada*); ii) “cold milk” (*leche fria*); iii) “hidden heat” (*calor recogido*); iv) “dull milk” (*empacho*); v) “choleric milk” (*cólera*); or vi) “spoiled milk” (*leche mala*).

Women indicated that regular breast milk transformed into “hot milk” if directly exposed to the sun and that the consumption of “hot milk” produced an odorous yellowish or whitish abundant diarrhoea and continuous vomiting. Mothers defined “cold milk” (or *chiriaiti*) as the opposite of “hot milk”, noting that breast milk became cold when mothers were in a cold environment or came into contact with a cold substance (e.g., water). This type of breast milk, they claimed, produced mucosal greenish diarrhoea and vomiting. Likewise, mothers linked the appearance of “hidden heat” to the time of year when temperature fluctuations were common. They described this ailment as a combination of “cold milk” and “hot milk”:

*“My first daughter was always ill and I did not know the reason. At the healthcare centre, the personnel told me it was diarrhoea. Thus, I gave her medicines but they never worked. Then, I went to the pharmacy and what they gave me did not work either. She was always ill. That is the reason why I tried a homemade treatment that people told me to use. It was to treat a disease between the “hot” and the “cold”. We call it “hidden heat”“* (Interview with Marina, August 8, 2016).

According to women, the fourth type of harmful breast milk, “dull milk”, was closely related to nutrition. A diet mainly composed of carbohydrates would make the breast milk thicker, generating stomach-aches and foamy stools in children. In contrast, “choleric milk” was thought to have a psychosomatic origin, as its appearance was associated with emotional distress:

*“Imagine that today I go to a place and people start saying horrible things about my family or me, like that my husband in an alcoholic. Then I get angry and I breastfeed my child. All that irritation and negative feelings pass to my child, who becomes sick. Children can die with this disease, because when they have “choleric milk” they vomit, have diarrhoea, their skin becomes pale, and their tummy is swollen”* (Interview with Lucrecia, July 7 2016).

Women described diarrhoea produced by “choleric milk” as yellowish and intense. In order to distinguish “hot milk” from “choleric milk” (both had a similar appearance), some women recommended rubbing an egg against the child’s extremities and then emptying it into a glass to see the “origin” of the disease. According to mothers, this practice was also used to identify other pathologies included in the local illness model, such as the “evil eye”. Mothers described “choleric milk” as particularly dangerous for children as it made them thinner and causes their nails to blacken. Mothers emphasised that “choleric milk’” was potentially fatal if treated at healthcare centres:

*“When children have “choleric milk” we do not go to healthcare centres. When it is too strong, doctors give injections that kill the child. At healthcare facilities, there is no medicine for “choleric milk”. What they use is worthless. However, when children take our herbs* [infusions] *or the anti-choleric syrup, they recover quickly” (Interview with Angela, July 19, 2016)*.

Finally, women asserted that milk got spoiled (“spoiled milk”) with a new pregnancy while still breastfeeding a toddler: “It is as if the other baby were jealous. This ailment delays pregnancy and makes children sick” (Interview with Teresa, August 15, 2016). “Spoiled milk” implied possible adverse pregnancy outcomes as well as diarrhoea and vomiting in the nursed child. Diarrhoea produced by “spoiled milk” would have a liquid texture and different colours (e.g., yellowish, greenish). Some stated that “spoiled milk” only appeared when the new child was of a different sex than the currently breastfed one.

Women also describe different treatments and methods of prevention for each local type of child diarrhoea. To prevent “hot milk”, mothers recommended expressing some breast milk before giving the breast to the child or avoiding breastfeeding under the sun. Mothers believed that diarrhoea produced by “hot milk” could only be treated with “cold” herbs. They also endorsed rubbing the child’s stomach and forehead with herbs such as the sacred leaf (*Piper auritum*) or chicory (*Chicorium intybus*). For preventing “cold milk”, mothers suggested drinking hot beverages before breastfeeding. Remedies included the use of “hot” herbs or homemade treatments such as rubbing children’s upper and lower extremities with hot water, massaging the chest with eucalyptus extract, or eating fried chicken fat. To treat “hidden heat”, mothers prepared beverages combining “cold” and “hot” ingredients. To prevent “dull milk”, mothers recommended consuming purgative herbs used for stomach cleansing (e.g., *sen* (*cassia acutifolia*) or olive oil). Treatments for “dull milk” included tipping children upside-down and patting them on the back to expel gases. Mothers indicated that the only way to prevent “choleric milk” was to avoid negative thoughts. Treatments included the use of “cold” herbs and the so-called anti-choleric syrups or pills. They were sold in pharmacies and grocery stores, and the price per dose varied between 0.1 and one Peruvian sol (0.03 – 0.30 $USD cents). Finally, mothers argued that the only treatment and solution for “spoiled milk” was breastfeeding cessation. As a general remark, women emphasised that none of these local types of child diarrhoea could not be treated with medical drugs. Table 2 describes the local explanatory model for child diarrhoea associated with breastfeeding practices.

**Table 2.** Local types of child diarrhoea associated to breastfeeding in rural Andean Peru

| Local name | Perceived cause | Symptoms | Diarrhoea-specific perceptions |  |  | Preventive practices and beliefs | Treatment preferences |
| --- | --- | --- | --- | --- | --- | --- | --- |
|  |  |  | Colour | Texture | Quantity |  |  |
| "Hot milk" | Hot | - Diarrhoea<br>- Fever<br>- Vomits | White or yellow | Pasty | Abundant | - Avoid breastfeeding in/after sun exposure | - "Cold" herbs<br>- Rubbings |
| "Cold milk"<sup>1</sup> | Cold | - Diarrhoea<br>- Vomits | Green | Mucous | Irregular | - Avoid breastfeeding in/after sun exposure<br>- Consumption of hot beverages | - "Hot" herbs<br>- Rubbings |
| "Hidden heat" | Hot and Cold | - Diarrhoea<br>- Vomits | - | - | - | - | - "Hot" herbs /<br>"Cold" herbs |
| "Dull milk" | Personal diet | - Abdominal pains<br>- Constipation<br>- Foaming stools | - | - | - | - Back slapping<br>- Stomach cleaning | - Back slapping<br>- Stomach cleaning |
| "Choleric milk" | Emotional status | - Diarrhoea<br>- Vomits | Yellow | Pasty | Abundant | - Avoid negative thoughts | - "Cold" herbs<br>- Rubbings<br>- Anti-choleric |
| "Spoiled milk" | New pregnancy | - Diarrhoea<br>- Vomits | Yellow or green | Liquid | - | - | - breastfeeding cessation |

<sup>1</sup> Also called *chiriaiiti*

Women participants associated the previously described set of locally defined types of child diarrhoea with two other ailments included in the local illness model: i) “infection” and ii) “fright sicknesses” (*susto*). “Infection” was a diarrhoea caused by bacteria, bugs (parasites and insects), and dirt that lived on the ground. Mothers described the stools associated with this ailment as having a pasty yellowish texture. To treat “infection” diarrhoea, mothers recommended the use of “cold” herbs and antibiotics such as Trimethoprim Sulfamethoxazole. Antibiotic treatment was though preferred due to the belief that children would heal faster. Mothers asserted breastfeeding was safe when a child had “infection” diarrhoea. For this reason, some also consumed antibiotics during their child’s diarrhoea, as they thought medicine would be transmitted through breast milk and accelerate the recovery. Overall, “infection” was the only child diarrhoea that women perceived as treatable with medicines.

“Fright sickness” (*pachachari*) was believed to have a psychosomatic origin: “It is when something scares our children. When they get the *pachachari*, they no longer eat nor sleep and they have strong diarrhoea” (Interview with Agueda, August 13, 2016). “Fright sickness” make children cry continuously and “makes them thinner”. Mothers described this type of diarrhoea as liquid and abundant (e.g., a texture similar to water) and with several colours (e.g., yellow, white, green). To treat “fright sickness”, families carried out rituals called *limpias*. These consisted in rubbing the child’s body with a special white stone (*lumbre*) at midnight on Tuesdays and Fridays repeating the ritual for three consecutive days. Some mothers indicated that it was also possible to perform the *limpia* ritual at noon. Then, the *lumbre* had to be thrown into a fire. When the fire went out, an image of the entity that had “scared” the child would appear in the ashes. Mothers emphasised the importance of hiding or removing these ashes as other people might use them to invoke the child’s soul and make black magic rituals. The *lumbre* stones were normally sold in pharmacies and grocery stores. Mothers did not mention breastfeeding was safe when a child had “fight sickness”.

### Integrating the local illness model into healthcare and service provision

The majority of health personnel recognised the existence of a local corpus of beliefs and interpretations concerning child diarrhoea. However, they stated not hearing often about these pathologies:

*“Today, we do not see too much of them* [local beliefs]. *Before, mothers said their children were malnourished because they had been scared. However, those beliefs are already modified. Maybe some families still believe those things, but we no longer see them” (Interview with the obstetrician Facunda, October 20, 2016)*.

Overall, interviews showed a lack of integration of local beliefs in routine medical care and counselling as well as a lack of consensus about how to accommodate them. The most experienced health personnel showed the greatest willingness to incorporate these local practices and beliefs. On the contrary, the less experienced healthcare workers believed that families should modify their attitudes. Some women participants indicated that negative attitudes of health personnel towards local beliefs stopped them from using healthcare facilities when their children had diarrhoea:

*“Last year, my child had fever and sores in his mouth, so I went to the healthcare centre. They gave him syrups but nothing happened. A neighbour told me to look for a special herb in order to prepare a beverage. My child drank it and recovered quickly. I think he had a stomach fever because his mouth was very hot. However, at the healthcare centre they told me that it was impossible, that it did not exist” (Interview with Paula, August 16, 2016)*.

Health personnel identified chronic lack of personnel and large staff turnover as the main factors responsible for worsening relationships with rural patient clients. They described conflict situations were common, especially when health trainees from urban Peru came to provinces to do their mandatory internship (The Rural and Urban Marginal Health Service (SERUMS) programme). Some health practitioners stated that, sometimes, participants of the SERUMS programme dismissed traditional concepts and beliefs during doctor-patient interactions:

*“They do not tell us about them anymore* [local beliefs] *because some personnel says “hey, this is not right”. Some personnel explain things kindly to mothers. Others, however, say directly “no” in front of them. That is what makes mothers not want to come back. Those are especially the new ones, the serumes* [personnel from the SERUM programme]. *They come from cities and have their own ideas. They do not care about what people think here. They come, do their service, and then leave again” (Interview with the nurse Patricia, November 2, 2016)*.

## Discussion

Our qualitative study describes cultural factors for breastfeeding cessation and their relationships to the local explanatory model of child diarrhoea in rural high-altitude Andean Peru. Mothers considered breastfeeding important for healthy fostering of new-born children. However, they also stated that breastfeeding might produce diarrhoea when in certain contexts and periods (e.g., while exposed to hot/cold environments). Mothers identified eight local types of diarrhoea, and associated six of them with breastfeeding. Among these types of child diarrhoea, mothers identified only one (“infection”) with hygiene and the germ disease concept, and perceived it as treatable through drug therapy. Interviews with mothers and local health personnel demonstrated a lack of integration of these beliefs within the local healthcare and service provision as well as a strong refusal among mothers to get treatment for all but one type of diarrhoea at healthcare centres.

### Socio-cultural dimensions of breastfeeding

Our study provides evidence that the “hot and cold” theory is culturally integrated into mothers’ perceptions of diarrhoea aetiology. The “hot and cold” theory describes the balance between physical and psychosocial states [10, 42, 43]. Previous research from other Andean settings present similar findings. In a study assessing indigenous and public health infant feeding recommendations in rural Southern Andean Peru, mothers argued that breast milk was a “cold” medicine used to cure “hot” ailments such as “hot” fevers, backache or eye infections. However, they also perceived that an unbalanced diet and exposure to various environmental factors (e.g., heat, cold, wind) contaminated breast milk. Expressing some breast milk before nursing was seen as a preventive practice, but mothers declared that extracted breast milk may cause illness when exposed to the sun [34]. In another study assessing health seeking behaviours in rural Andean Peru and Bolivia, mothers reported that diarrhoeas could be caused by breast milk from a mother experiencing emotional distress (*cólera*) [35]. Finally, a study conducted in the Southern Puno region (Peru) described how Andean women had to dress warmly while breastfeeding to prevent the transmission of “cold”-related diarrhoeas to their children [44].

Similar perceptions are observed in other LMIC. In India, Tanzania and Thailand, mothers believe their emotional and health state is transmitted through breast milk [16, 17, 45].

Evidence suggests that breastfeeding is an intersubjective behaviour that is articulated differently between cultural, social, economic, biological, psychological, symbolic, and political dimensions [46-48]. In all societies, breastfeeding is used as a means for managing emotions, reinforcing social norms, defining cultural images and identities, and strengthening mother-child relationships [49-52]. For example, in rural Mexico, breastfeeding becomes less accepted (and even reproached) when the child had more than a year or starts growing teeth. Moreover, women perceived that breastfeeding in public is sometimes depicted as a practice for poor, rural and indigenous women [53].

### Peer-counselling breastfeeding approaches

All mothers in our study received breastfeeding counselling at healthcare facilities. Yet, we found that cultural perceptions regarding breastfeeding prevailed and breastfeeding cessation was common. A potential explanation is the approach used to promote breastfeeding. Some authors argue that promoting specific ideas and principles regarding breastfeeding practices (e.g., WHO recommendations) without considering the local context may generate blurred interpretations and decrease the effectiveness of breastfeeding interventions [47, 48, 54, 55]. In this regard, peer-counselling breastfeeding programmes using trained women living in the communities proved to be effective to improve rates of breastfeeding initiation, duration, and exclusivity and reduce child diarrhoea incidence in rural settings [56, 57]. In addition, they are cost-effective [58, 59] and their effects can be sustained over time [60]. The main factor behind the success of peer-counselling breastfeeding programmes is community engagement, as mothers highly appreciate to have someone from their own community to help them with their breastfeeding problems [61, 62].

### Health-seeking behaviours for treating child diarrhoea

We found that mothers in our study did not consider breastfeeding useful against local types of child diarrhoea; therefore, other treatment options (herbal medicine) were used. This contradicts the biomedical message that breastfeeding is effective for treating child diarrhoea. Overall, mothers noted that breastfeeding was only useful for treating one diarrhoea linked to hygiene and the germ disease concept (“infection”). They also declared using antibiotics to treat this diarrhoea due to the perception that they cured diarrhoea faster. Ideas about the efficacy and effectiveness of medicines have marked cultural dimensions. In Southeast Asia and Central and South America, some people doubt the efficacy of oral re-hydration solution for treating diarrhoea because its appearance does not resemble medicine [12, 63, 64]. Studies highlight antibiotics are commonly used (with and without medical prescription) for treating non-antibiotic-associated child diarrhoea in Peru and other LMIC [65, 66]. This practice is not recommended by the WHO and has negative effects on anti-microbial resistance and the infant gut microbial development [67]. In our setting, women declared using antibiotics to treat “infection” diarrhoea and consumed antibiotics at the same time. They believed that healing effects of antibiotics were transmitted to their children throughout breast milk, reducing recovering times. Similar behaviours are reported in other LMIC such as Thailand [16].

### Local beliefs and healthcare and service provision

The incorporation of local beliefs and traditional medicine practices within the biomedical system is critical for delivering adequate quality of care and improve health outcomes [68-70]. We found a poor level of integration of the local explanatory model of child diarrhoea within the healthcare and service provision in our setting. In addition, we found that mothers had negative attitudes towards health personnel, which may engender important harmful effects such as the inappropriate management of bacterial diarrhoeas and the possible aggravation of symptoms. Cultural competence education for health professional gained relevance in recent years [68], and different studies emphasise that South American illness explanatory models can be feasibly incorporated into local healthcare and service provision [35, 71-74]. However, systematic reviews of interventions applying cultural competence education approaches to date only found moderate improvements in health provider outcomes and healthcare access and weak improvements in patient-provider relationships. The heterogeneity of intervention strategies, measures and outcomes are the main limitations described [75, 76].

### Limitations

Our study is not without limitations. The qualitative design does not allow making any inference to what extent these perceived causes are prevalent and how mothers manage child diarrhoea episodes. In addition, we did not investigate the impact of social networks in the preservation and diffusion of these beliefs, although some of the interview extracts showed that these factors may be important. Finally, the enrolment of participants with similar socio-economic and educational background (derived from IHIP-2 inclusion criteria) made it difficult to identify alternative breastfeeding discourses. We cannot state that socio-cultural factors for breastfeeding cessation are predominant in the study setting. Future research would need to compare perceptions of mothers from different socio-economic backgrounds, education levels, and occupations.

### Practical applications of the results

Despite these limitations, we believe our study is especially interesting for researches, health professionals and policymakers, as it describes practices and beliefs that could be incorporated into programmes that are currently active in rural Andean Peru. The Peruvian national ECD programme (PNCM) was launched in 2012, to improve the cognitive, social, and emotional development of infants <36 months living in poverty. In rural areas, the PNCM provides home-visiting interventions consisting in weekly play-oriented activities and monthly communal meetings carried out by trained women living in the communities (mother facilitators -MFs). In 2018, a new decree fostered the promotion of breastfeeding practices in the PNCM by encouraging breastfeeding practices and counselling pregnant women for immediate breastfeeding after childbirth. The decree specifically focused on technical aspects of breastfeeding (e.g., position, timing), but other structural and socio-cultural dimensions were not discussed. Despite the breastfeeding component of the PNCM does not currently follow a peer-counselling approach, the structure of PNCM’s weekly visits and the role of MFs could facilitate its inclusion as local populations are already involved in regular PNCM activities and rely on MFs.

In the same vein, the incorporation of cultural breastfeeding and child diarrhoea interpretations into clinical research can be useful to provide alternative explanations to unforeseen results. Recent large-scale randomised controlled trials found that single and combined water and sanitation interventions had no impact in reducing child diarrhoea despite robust research designs [77, 79]. In addition, other studies were not able to explain why Peruvian lactating women using traditional contraceptive methods (e.g., periodic abstinence or withdrawal) breastfed longer than those using oral contraceptives or injections [80]. Applying the connections between local explanatory models, breastfeeding, and child diarrhoea in similar investigations may help to interpret these uncertainties.

## Conclusion

Women with low socio-economic and education backgrounds living in rural high-altitude Andean Peru identified cultural beliefs for breastfeeding cessation that were linked to child diarrhoea. These perceptions were not integrated within the local healthcare and service provision, resulting in negative attitudes towards health personnel. Future breastfeeding interventions in rural Andean Peru should promote peer-counselling approaches. Introducing mother’s concerns and cultural breastfeeding interpretations into the National programme on early child development could be a feasible and cost-effective solution. Additional research on cultural factors for breastfeeding cessation are necessary to promote equity in health.

## Data Availability

The datasets used and/or analysed during the current study are available from the corresponding author on reasonable request.

## List of abbreviations

COREQ: Consolidated Criteria for Reporting Qualitative Research
ECD: early child development
IHIP-2: integrated home-based intervention package trial 2
LMIC: low- and middle-income countries
PNCM: Peruvian National programme on early child development
UPCH: Cayetano Heredia University
WHO: World Health Organization

## Declarations

### Ethics approval and consent to participate

Ethical approval for IHIP-2 (trial registry: ISRCTN26548981) was obtained from the ethics commissions of the Universidad Peruana Cayetano Heredia (UPCH) and the Cajamarca Regional Health Authority. Additional ethical approval for our study was obtained from the ethics commission of the UPCH (Ref. 61065). Women participants signed an informed consent form, and health personnel provided tape-recorded consent. We deleted community designations and participant names to ensure compliance with ethical principles of research. The information obtained from the study was treated confidentially.

## Consent for publication

Not applicable.

## Author’s contributions

DM and SH obtained the funding; NN and DM designed the study; NN collected data; NN and JM analysed and interpreted the data; NN wrote the first draft manuscript; JW, DM, NN, SH and JM performed critical revisions of the manuscript and contributed to the writing; SH and DM provided administrative, technical and material support; SH, NN, and DM, coordinated and supervised the study.

## Acknowledgments

The authors would like to express their appreciation to study women and health personnel of the RedSalud-IV and RedSalud-V for their kind participation. We also express our gratitude Mrs. Angelica Fernandez and Ms. Maria Luisa Huaylinos, field coordinators of the IHIP-2 trial. Finally, we thank Marie Reinholdt for proofreading the manuscript.

## References

1. Troeger C, Blacker BF, Khalil IA, Rao PC, Cao S, Zimsen SRM et al. Estimates of the global, regional, and national morbidity, mortality, and aetiologies of diarrhoea in 195 countries: a systematic analysis for the Global Burden of Disease Study 2016. The Lancet Infectious Diseases. 2018;18(11):1211–28. doi:10.1016/S1473-3099(18)30362-1.

2. Brown KH. Diarrhea and Malnutrition. The Journal of Nutrition. 2003;133(1):328S–32S. doi:10.1093/jn/133.1.328S.

3. Checkley W, Buckley G, Gilman RH, Assis AM, Guerrant RL, Morris SS et al. Multi-country analysis of the effects of diarrhoea on childhood stunting. International Journal of Epidemiology. 2008;37(4):816–30. doi:10.1093/ije/dyn099.

4. Warren-Gash C, Fragaszy E, Hayward AC. Hand hygiene to reduce community transmission of influenza and acute respiratory tract infection: a systematic review. Influenza and Other Respiratory Viruses. 2013;7(5):738–49. doi:10.1111/irv.12015.

5. Black RE, Allen LH, Bhutta ZA, Caulfield LE, de Onis M, Ezzati M et al. Maternal and child undernutrition: global and regional exposures and health consequences. The Lancet. 2008;371(9608):243–60. doi:10.1016/S0140-6736(07)61690-0.

6. Guerrant RL, DeBoer MD, Moore SR, Scharf RJ, Lima AA. The impoverished gut--a triple burden of diarrhoea, stunting and chronic disease. Nature reviews Gastroenterology & hepatology. 2013;10(4):220–9. doi:10.1038/nrgastro.2012.239.

7. Scharf RJ, Deboer MD, Guerrant RL. Recent advances in understanding the long-term sequelae of childhood infectious diarrhea. Current Infectious Disease Report. 2014;16(6):408-. doi:10.1007/s11908-014-0408-y.

8. Victora CG, Bahl R, Barros AJ, Franca GV, Horton S, Krasevec J et al. Breastfeeding in the 21st century: epidemiology, mechanisms, and lifelong effect. The Lancet. 2016;387(10017):475–90. doi:10.1016/s0140-6736(15)01024-7.

9. Kleinman A. Patients and Healers in the Context of Culture. An Exploration of the Borderland between Anthropology, Medicine, and Psychiatry. Berkeley: University of California Press; 1980.

10. Weiss MG. Cultural models of diarrheal illness: Conceptual framework and review. Social Science & Medicine. 1988;27(1):5–16. doi:10.1016/0277-9536(88)90159-1.

11. Nuño N, Muela J, Hausmann-Muela S, Cevallos M, Hartinger S, Christen A et al. The Meanings of Water: Socio-Cultural Perceptions of Solar Disinfected (SODIS) Drinking Water in Bolivia and Implications for its Uptake. Water. 2020;12(2). doi:10.3390/w12020442.

12. Vázquez ML, Mosquera M, Kroeger A. People’s concepts on diarrhea and dehydration in Nicaragua: the difficulty of the intercultural dialogue. Revista Brasileira de Saúde Materno Infantil. 2002;2:223–37. doi:10.1590/S1519-38292002000300003.

13. Pitts M, McMaster J, Hartmann T, Mausezahl D. Lay beliefs about diarrhoeal diseases: Their role in health education in a developing country. Social Science & Medicine. 1996;43(8):1223–8. doi:10.1016/0277-9536(95)00430-0.

14. Nkwi PN. Perceptions and treatment of diarrhoeal diseases in Cameroon. Journal of Diarrhoeal Diseases Research. 1994;12(1):35–41.

15. Rheinländer T, Samuelsen H, Dalsgaard A, Konradsen F. Perspectives on child diarrhoea management and health service use among ethnic minority caregivers in Vietnam. BMC Public Health. 2011;11(1):690. doi:10.1186/1471-2458-11-690.

16. Pylypa J. Elder Authority and the Situational Diagnosis of Diarrheal Disease as Normal Infant Development in Northeast Thailand. Qualitative Health Research. 2009;19(7):965–75. doi:10.1177/1049732309338867.

17. Mabilia M. The cultural context of childhood diarrhoea among Gogo infants. Anthropology & Medicine. 2000;7(2):191–208. doi:10.1080/713650590.

18. Ene-Obong HN, Iroegbu CU, Uwaegbute AC. Perceived causes and management of diarrhoea in young children by market women in Enugu State, Nigeria. Journal of Health, Population, and Nutrition. 2000;18(2):97–102.

19. Mull DS. Mother’s milk and pseudoscientific breastmilk testing in Pakistan. Social Science & Medicine. 1992;34(11):1277–90. doi:10.1016/0277-9536(92)90320-p.

20. Nichter M, Nichter M. Anthropology and International Health. Asian Case Studies. The Netherlands: Gordon and Breach; 1996.

21. Tromp I, Kiefte-de Jong J, Raat H, Jaddoe V, Franco O, Hofman A et al. Breastfeeding and the risk of respiratory tract infections after infancy: The Generation R Study. PLoS One. 2017;12(2):e0172763. doi:10.1371/journal.pone.0172763.

22. Rollins NC, Bhandari N, Hajeebhoy N, Horton S, Lutter CK, Martines JC et al. Why invest, and what it will take to improve breastfeeding practices? The Lancet. 2016;387(10017):491–504. doi:10.1016/S0140-6736(15)01044-2.

23. Ladomenou F, Moschandreas J, Kafatos A, Tselentis Y, Galanakis E. Protective effect of exclusive breastfeeding against infections during infancy: a prospective study. Archives of Disease in Childhood. 2010;95(12):1004. doi:10.1136/adc.2009.169912.

24. Pecoraro L, Agostoni C, Pepaj O, Pietrobelli A. Behind human milk and breastfeeding: not only food. International Journal of Food Sciences and Nutrition. 2018;69(6):641–6. doi:10.1080/09637486.2017.1416459.

25. United Nations Children’s Fund. Infant and Young Child Feeding. New York: UNICEF 2011.

26. Forster DA, McLachlan HL, Lumley J. Factors associated with breastfeeding at six months postpartum in a group of Australian women. International Breastfeeding Journal. 2006;1:18-. doi:10.1186/1746-4358-1-18.

27. Kelly YJ, Watt RG. Breast-feeding initiation and exclusive duration at 6 months by social class--results from the Millennium Cohort Study. Public Health Nutrition. 2005;8(4):417–21. doi:10.1079/phn2004702.

28. Yeneabat T, Belachew T, Haile M. Determinants of cessation of exclusive breastfeeding in Ankesha Guagusa Woreda, Awi Zone, Northwest Ethiopia: a cross-sectional study. BMC Pregnancy Childbirth. 2014;14:262. doi:10.1186/1471-2393-14-262.

29. Su L, Chong Y, Chan Y, Chan Y, Fok D, Tun K et al. Antenatal education and postnatal support strategies for improving rates of exclusive breast feeding: randomised controlled trial. BMJ. 2007;335(7620):596. doi:10.1136/bmj.39279.656343.55.

30. Wallenborn JT, Ihongbe T, Rozario S, Masho SW. Knowledge of Breastfeeding Recommendations and Breastfeeding Duration: A Survival Analysis on Infant Feeding Practices II. Breastfeed Medicine. 2017;12:156–62. doi:10.1089/bfm.2016.0170.

31. Araujo MC, López Bóo F, Puyana JM. Overview of Early Childhood Development Services in Latin America and the Caribbean: Inter-American Development Bank2013.

32. Instituto Nacional de Estadística e Informática. Encuesta Demográficay de Salud Familiar-ENDES. Lima: INEI; 2015.

33. Froemming S. Traditional use of the Andean flicker (Colaptes rupicola) as a galactagogue in the Peruvian Andes. Journal of Ethnobiology and Ethnomedicine. 2006;2(1):23. doi:10.1186/1746-4269-2-23.

34. Monteban M, Yucra Velasquez V, Yucra Velasquez B. Comparing Indigenous and public health infant feeding recommendations in Peru: opportunities for optimizing intercultural health policies. Journal of Ethnobiology and Ethnomedicine. 2018;14(1):69. doi:10.1186/s13002-018-0271-2.

35. Mathez-Stiefel S, Vandebroek I, Rist S. Can Andean medicine coexist with biomedical healthcare? A comparison of two rural communities in Peru and Bolivia. Journal of Ethnobiology and Ethnomedicine. 2012;8(1):26. doi:10.1186/1746-4269-8-26.

36. Escobar GJ, Salazar E, Chuy M. Beliefs regarding the etiology and treatment of infantile diarrhea in Lima, Peru. Social Science & Medicine. 1983;17(17):1257–69. doi:10.1016/0277-9536(83)90018-7.

37. Hartinger SM, Nuño N, Verastegui H, Ortiz M, Mäusezahl D. A factorial randomised controlled trial to combine early child development and environmental interventions to reduce the negative effects of poverty on child health and development: rationale, trial design and baseline findings. BMC Medical Research Methodology. 2020;20(73). doi:10.1186/s12874-020-00950-y.

38. Tong, A, Sainsbury P, Craig J. Consolidated criteria for reporting qualitative research (COREQ): a 31-item checklist for interviews and focus groups. International Journal for Quality in Health Care. 2007; 19(6): 349-357. doi.org/10.1093/intqhc/mzm042.

39. Bryman A. Social Research Methods. Oxford: Oxford University Press; 2008.

40. Green EC, Jurg A, Djedje A. The Snake in the Stomach: Child Diarrhea in Central Mozambique. Medical Anthropology Quarterly. 1994;8(1):4–24. doi:10.1525/maq.1994.8.1.02a00020.

41. Briones K. ‘How Many Rooms Are There in Your House?’ Constructing the Young Lives Wealth Index. Oxford: UKAid 2017.

42. Messer E. Hot-cold classification: Theoretical and practical implications of a Mexican study. Social Science & Medicine. 1981;15(2):133–45. doi:10.1016/0160-7987(81)90036-3.

43. Mazess RB. Hot-cold food beliefs among Andean peasants. Journal of the American Dietetic Association. 1968;53(2):109–13.

44. Larme AC. Environment, vulnerability, and gender in Andean ethnomedicine. Social Science & Medicine. 1998;47(8):1005–15. doi:10.1016/S0277-9536(98)00162-2.

45. Reissland N, Burghart R. The quality of a mother’s milk and the health of her child: beliefs and practices of the women of Mithila. Social Science & Medicine. 1988;27(5):461–9. doi:10.1016/0277-9536(88)90369-3.

46. Esteban ML. La maternidad como cultura. Algunas cuestiones sobre lactancia materna y cuidado infantil. In: Pedriguero E, Comelles JM, editors. Medicina y cultura: estudios entre la medicina y la antropología. Barcelona: Bellaterra; 2000.

47. Thorley V. Is breastfeeding ‘normal’? Using the right language for breastfeeding. Midwifery. 2019;69:39–44. doi:10.1016/j.midw.2018.10.015.

48. Fouts HN, Hewlett BS, Lamb ME. A Biocultural Approach to Breastfeeding Interactions in Central Africa. American Anthropologist. 2012;114(1):123–36. doi:10.1111/j.1548-1433.2011.01401.x.

49. Clarke M. The modernity of milk kinship. Social Anthropology. 2007;15(3):287–304. doi:10.1111/j.0964-0282.2007.00022.x.

50. Shaw R. The virtues of cross-nursing and the ‘yuk factor. Australian Feminist Studies. 2004;19(45):287–99. doi:10.1080/0816464042000278972.

51. Dettwyler KA. When to wean: biological versus cultural perspectives. Clinical Obstetrics and Gynecology. 2004;47(3):712–23. doi:10.1097/01.grf.0000137217.97573.01.

52. Dodgson J, Struthers R. Indigenous Women’s Voices: Marginalization and Health. Journal of Transcultural Nursing. 2005;16:339–45. doi:10.1177/1043659605278942.

53. Swigart TM, Bonvecchio A, Théodore FL, Zamudio-Haas S, Villanueva-Borbolla MA, Thrasher JF. Breastfeeding practices, beliefs, and social norms in low-resource communities in Mexico: Insights for how to improve future promotion strategies. PLoS One. 2017;12(7):e0180185. doi:10.1371/journal.pone.0180185.

54. Fewtrell MS, Mohd S. NH, Wells JCK. ‘Optimising’ breastfeeding: what can we learn from evolutionary, comparative and anthropological aspects of lactation? BMC Medicine. 2020;18(1):4. doi:10.1186/s12916-019-1473-8.

55. Olza I, Ruiz-Berdún D, Villarmea S. La culpa de las madres. Promover la lactancia materna sin presionar a las mujeres. Dilemata Revista Internacional de Éticas Aplicadas. 2017;25:217–25.

56. Chapman DJ, Morel K, Anderson AK, Damio G, Pérez-Escamilla R. Breastfeeding peer counseling: from efficacy through scale-up. Journal of Human Lactation. 2010;26(3):314–26. doi:10.1177/0890334410369481.

57. Shakya P, Kunieda MK, Koyama M, Rai SS, Miyaguchi M, Dhakal S et al. Effectiveness of community-based peer support for mothers to improve their breastfeeding practices: A systematic review and meta-analysis. PLoS One. 2017;12(5):e0177434–e. doi:10.1371/journal.pone.0177434.

58. Chola L, Fadnes LT, Engebretsen IM, Nkonki L, Nankabirwa V, Sommerfelt H et al. Cost-Effectiveness of Peer Counselling for the Promotion of Exclusive Breastfeeding in Uganda. PLoS One. 2015;10(11):e0142718. doi:10.1371/journal.pone.0142718.

59. Nkonki L, Tugendhaft A, Hofman K. A systematic review of economic evaluations of CHW interventions aimed at improving child health outcomes. Human Resources for Health. 2017;15(1):19. doi:10.1186/s12960-017-0192-5.

60. Haider R, Saha KK. Breastfeeding and infant growth outcomes in the context of intensive peer counselling support in two communities in Bangladesh. International Breastfeeding Journal. 2016;11(1):18. doi:10.1186/s13006-016-0077-6.

61. Nankunda J, Tumwine JK, Soltvedt A, Semiyaga N, Ndeezi G, Tylleskär T. Community based peer counsellors for support of exclusive breastfeeding: experiences from rural Uganda. International Breastfeeding Journal. 2006;1:19. doi:10.1186/1746-4358-1-19.

62. Rujumba J, Ndeezi G, Nankabirwa V, Kwagala M, Mukochi M, Diallo AH et al. “If I have money, I cannot allow my baby to breastfeed only …” barriers and facilitators to scale-up of peer counselling for exclusive breastfeeding in Uganda. International Breastfeeding Journal. 2020;15(1):43-. doi:10.1186/s13006-020-00287-8.

63. White SR, Van der Geest S, Hardon A. Social Lives of Medicines. Cambridge: Cambridge University Press; 2002.

64. Bentley ME. The household management of childhood diarrhea in rural north India. Social Science & Medicine. 1988;27(1):75–85. doi:10.1016/0277-9536(88)90165-7.

65. Ecker L, Ochoa TJ, Vargas M, Del Valle LJ, Ruiz J. Factors affecting caregivers’ use of antibiotics available without a prescription in Peru. Pediatrics. 2013;131(6):e1771–9. doi:10.1542/peds.2012-1970.

66. Carter E, Bryce J, Perin J, Newby H. Harmful practices in the management of childhood diarrhea in low-and middle-income countries: a systematic review. BMC Public Health. 2015;15(1):788. doi:10.1186/s12889-015-2127-1.

67. World Health Organization. The Treatment of Diarrhoea. A manual for physicians and other senior health workers. Geneva: World Health Organization; 2005.

68. Saha S, Beach MC, Cooper LA. Patient centeredness, cultural competence and healthcare quality. Journal of the National Medical Association 2008;100(11):1275–85. doi:10.1016/s0027-9684(15)31505-4.

69. Kayne SB. Traditional Medicine. A Global Perspective. London: Pharmaceutical Press; 2010.

70. Kahissay MH, Fenta TG, Boon H. Beliefs and perception of ill-health causation: a socio-cultural qualitative study in rural North-Eastern Ethiopia. BMC Public Health. 2017;17(1):124. doi:10.1186/s12889-017-4052-y.

71. Hudelson P, Huanca T, Charaly D, Cirpa V. Ethnographic studies of ARI in Bolivia and their use by the national ARI programme. Social Science & Medicine. 1995;41(12):1677–83. doi:10.1016/0277-9536(95)00128-T.

72. Kendall C, Foote D, Martorell R. Ethnomedicine and oral rehydration therapy: A case study of ethnomedical investigation and program planning. Social Science & Medicine. 1984;19(3):253–60. doi:10.1016/0277-9536(84)90216-8.

73. Scrimshaw SCM, Hurtado E. Anthropological involvement in the Central American diarrheal disease control project. Social Science & Medicine. 1988;27(1):97–105. doi:10.1016/0277-9536(88)90167-0.

74. Bentley ME, Pelto GH, Straus WL, Schumann DA, Adegbola C, de la Pena E et al. Rapid ethnographic assessment: Applications in a diarrhea management program. Social Science & Medicine. 1988;27(1):107–16. doi:10.1016/0277-9536(88)90168-2.

75. Truong M, Paradies Y, Priest N. Interventions to improve cultural competency in healthcare: a systematic review of reviews. BMC Health Services Research. 2014;14:99. doi:10.1186/1472-6963-14-99.

76. Jongen C, McCalman J, Bainbridge R. Health workforce cultural competency interventions: a systematic scoping review. BMC Health Services Research. 2018;18(1):232-. doi:10.1186/s12913-018-3001-5.

77. Null C, Stewart CP, Pickering AJ, Dentz HN, Arnold BF, Arnold CD et al. Effects of water quality, sanitation, handwashing, and nutritional interventions on diarrhoea and child growth in rural Kenya: a cluster-randomised controlled trial. The Lancet Global Health. 2018;6(3):e316–e29. doi:10.1016/s2214-109x(18)30005-6.

78. Humphrey JH, Mbuya MNN, Ntozini R, Moulton LH, Stoltzfus RJ, Tavengwa NV et al. Independent and combined effects of improved water, sanitation, and hygiene, and improved complementary feeding, on child stunting and anaemia in rural Zimbabwe: a cluster-randomised trial. The Lancet Global Health. 2019;7(1):e132–e47. doi:10.1016/s2214-109x(18)30374-7.

79. Arnold BF, Khush RS, Ramaswamy P, London AG, Rajkumar P, Ramaprabha P et al. Causal inference methods to study nonrandomized, preexisting development interventions. Proceedings of the National Academy of Sciences. 2010;107(52):22605–10. doi:10.1073/pnas.1008944107.

80. Rice S, Coombs D, Fish L, Leeper J. Breast-feeding and contraception in Peru. Journal of Health, Population, and Nutrition. 2002;20(1):51–8.

